# Circulating Tumor DNA in Patients with Early-Onset Systemic Sclerosis and Dermatomyositis

**DOI:** 10.64898/2026.11.06.26364657

**Authors:** Christopher A. Mecoli, Yuxuan Wang, Ji Soo Kim, XingYao Wang, Adrianne Woods, Christopher Douville, Sam D. Curtis, Maria Popoli, Janine Ptak, Lisa Dobbyn, Natalie Silliman, Matthew B. Smith, Lauren Fisher, Gwen Leatherman, Laura K. Hummers, Lisa Christopher-Stine, Julie J. Paik, Jemima Albayda, Fredrick M. Wigley, Antony Rosen, Livia Casciola-Rosen, Chetan Bettegowda, Nickolas Papadopoulos, Kenneth W. Kinzler, Bert Vogelstein, Ami A. Shah

**Affiliations:** Division of Rheumatology, Department of Medicine, The Johns Hopkins University School of Medicine; Department of Oncology, the Sidney Kimmel Comprehensive Cancer Center, Johns Hopkins University School of Medicine; The Howard Hughes Medical Institute

## Abstract

**Background:** Systemic sclerosis (SSc) and dermatomyositis (DM) are autoimmune diseases that have been proposed as models of cancer immunoediting. Autoantibody-defined subsets show divergent cancer risks, yet malignancy often remains clinically occult. We tested whether circulating tumor DNA (ctDNA) can reveal subclinical cancer in new-onset SSc and DM.

**Methods:** We prospectively recruited adults with recent SSc or DM onset. Plasma ctDNA was assessed using a targeted next-generation sequencing panel covering commonly mutated cancer genes. Patients were followed for 18–24 months on average. ctDNA positivity was correlated with subsequent cancer diagnoses.

**Findings:** Among 134 patients (69 SSc, 65 DM), cancer was diagnosed in six (4·5%; three SSc, three DM). ctDNA was detected in four of these six patients (67%) a median of 13 months (range 1·4–17) before clinical diagnosis; in three of four patients, mutations detected in plasma matched those subsequently identified in the same patients’ primary tumors. ctDNA was also detected in 16 of 128 patients without a cancer diagnosis. ctDNA positivity conferred an 11·4-fold higher risk of malignancy relative to ctDNA negativity and yielded a negative predictive value of 98% and a positive predictive value of 20% for cancer within the observation window.

**Interpretation:** ctDNA testing identified most cancers that emerged within 18 months in new-onset SSc and DM and stratified near-term cancer risk. These findings support larger, interventional trials to determine whether ctDNA-guided surveillance improves timeliness of cancer detection, optimizes imaging and biopsy strategies, and enhances outcomes in patients with SSc and DM.

**Funding:** NIH, Scleroderma Research Foundation, Stabler Foundation, Chresanthe Staurulakis Memorial Fund, Sara and Alex Othon Research Fund, Bechtle Precision Medicine Fund for Scleroderma, Zhang Discovery Fund, Dr. Peter Buck Fund, Jerome L. Greene Foundation, Lustgarten Foundation for Pancreatic Cancer Research, Sol Goldman Sequencing Facility, Virginia and D.K. Ludwig Fund for Cancer Research.

**RESEARCH IN CONTEXT:** **1. Evidence before this study:** Patients with new onset systemic sclerosis (SSc) and dermatomyositis (DM) have an increased risk of contemporaneous cancer in distinct autoantibody subgroups, and mechanistic studies suggest that these patients may have cancer-induced autoimmunity. The optimal approach to cancer detection, and whether subclinical malignancy may be present in patients with new onset rheumatic disease, remains unknown.

**2. Added value of this study:** We show that circulating tumor DNA (ctDNA) is present in ∼15% of patients with new onset SSc and DM. Patients with ctDNA have a higher risk of subsequent malignancy, with data suggesting that it can detect cancer earlier than conventional methods in this patient population.

**3. Implications of all the available evidence:** These findings suggest that subclinical malignancy may be present in a subset of patients with new onset SSc and DM and that ctDNA may be a valuable tool to risk stratify patients for cancer.

## INTRODUCTION

The relationship between cancer and autoimmunity has attracted increasing attention over the past two decades. The concept of cancer immunoediting, encompassing the processes of cancer elimination, equilibrium, and escape, has been central to elucidating interactions between the immune system and cancer. This framework has advanced understanding of both effective immune surveillance and immune dysregulation, including the emergence of autoimmune phenomena. Systemic sclerosis (SSc) and dermatomyositis (DM) are two autoimmune rheumatic diseases characterized by a substantial association with cancer in patient subgroups defined by distinct autoantibodies (1, 2). For example, patients with anti-RNA polymerase III (POLR3)-positive SSc and anti-TIF1γ-positive DM have an increased risk of cancer within 3 years of rheumatic disease onset. Prior research has identified genetic alterations, including somatic mutations and loss of heterozygosity in the *POLR3A* and *TRIM33* genes which encode POLR3 and TIF1γ, respectively, in tumors from patients with anti-POLR3-positive SSc and anti-TIF1γ-positive DM (3–4). Furthermore, in patients with SSc, it has been shown that there are mutation-specific T cell and cross-reactive autoantibody responses to both mutated and wildtype forms of these antigens. Collectively, these findings suggest that an immune response initiated by somatic mutations in emerging tumors may inadvertently result in the targeting of healthy tissues, thereby triggering autoimmunity (5). Over the last decade, this emerging model has been supported by additional studies (6–8), reinforcing the concept that the immune system’s responses to tumors can have far-reaching consequences, in some cases leading to autoimmune disease.

Multiple studies have shown that only a subset of anti-POLR3-positive SSc and anti-TIF1γ-positive DM patients develop clinically detectable cancer (1–2, 6–8). One explanation for this heterogeneity is that the immune responses to POLR3 and TIF1γ might be induced by multiple mechanisms, with cancer being only one of these. Alternately, it is possible that cancer is a driver of these autoimmune responses in *all* patients within these serologically-defined subsets. The second explanation posits that in some patients, the immune system may successfully hold a nascent tumor in equilibrium preventing its progression to a clinically manifest cancer, perhaps even destroying the cancer. Support for this idea comes from studies showing that patients with specific SSc and DM autoantibodies have a lower than expected risk of cancer (1, 2). In particular, patients with SSc and anti-POLR3 plus anti-RNA polymerase 1 (POLR1) or anti-centromere (CENP) antibodies have a lower cancer risk relative to the general population. Additionally, breast cancer patients without SSc who have anti-CENP antibodies have improved cancer survival (9). In other patients, the immune response is not strong enough to control tumor progression, and clinically manifest cancer. Note that in both of these hypothetical scenarios, the cancer *precedes* the autoimmune disease.

Until recently, technological limitations restricted the detection and monitoring of early malignancies to modalities such as whole body [^18^F] fluorodeoxyglucose positron emission tomography/computed tomography (FDG-PET/CT) imaging and flow cytometry). The advent of liquid biopsy technologies, which enable detection of circulating tumor DNA (ctDNA) (10–12), has broadened this potential. In the current study, we aimed to determine whether ctDNA could be detected in patients with early onset SSc and DM prior to clinical presentation of cancer.

## METHODS

### Study Design and Participants

We conducted a prospective observational cohort study across two disease-specific centers, recruiting 134 patients from the Johns Hopkins Division of Rheumatology Scleroderma and Myositis Centers. Eligible patients had either SSc meeting 2013 ACR/EULAR criteria or DM meeting 2017 ACR/EULAR criteria, age ≥ 18 years, and a disease duration of less than 3 years. No history of cancer within 3 years of disease onset was allowed, excluding non-melanoma skin cancer. Patient recruitment began in November 2018, initially targeting individuals with SSc who were anti-POLR3 positive. In 2021, the criteria were expanded to include all patients with SSc, regardless of autoantibody status, to enable comparisons across serologic subgroups. Eligible DM patients were recruited consecutively for enrollment starting in May of 2022. The study protocols were approved by the Johns Hopkins Institutional Review Board (NA_00036834 and IRB00235256). Written informed consent was obtained from participants prior to study participation. All patients were followed longitudinally at the Johns Hopkins Scleroderma and Myositis Centers.

### Procedures

All 134 patients had at least one blood sample collected. Of these, 73 had a second sample collected at least 3 months after the first. Plasma from 50 mL of blood collected in Streck tubes was purified using BioChain cfDNA Extraction Kit (BioChain, cat #K5011610) using the manufacturer’s protocol. cfDNA was quantified using either electrophoresis (TapeStation cfDNA, Agilent catalog #5067-5630) or via qPCR using Sso Advanced SYBR Green Supermix (BioRad Cat #1725271) as directed by the manufacturer and employing the following primers: 5’-CACACAGGAAACAGCTATGACCATGGGTAACAGCTTTATCTATTGACATTATGC-3’ and 5’-CGACGTAAAACGACGGCCAGTNNNNNNNNNNNNNNAAACTTCATGCTTCATCTAGT CAGC-3’. National Institute of Standards and Technology (NIST) human DNA quantification standard NIST SRM 2372a, diluted to 1 ng/ml, served as the reference standard. 2·5 μL of cfDNA or NIST 2372a DNA was added to 97·5 μL of 1:1,000 SYBR Green I diluted in 1X PBS.

Amplification and fluorescence detection conditions were as follows: one cycle of 98°C for 120 seconds and then 30 cycles of 98°C for 10 seconds, 57°C for 120 seconds, and 72°C for 120 seconds. Leukocyte DNA was purified from whole blood and sheared to an average size of ∼200bp using a Hielscher UIP400MTP sonicator prior to library preparation as previously described (13). DNA was purified from macro-dissected tumors as previously described (14).

### Library preparation and targeted sequencing

SaferSeqS libraries were made from plasma and leukocytes as previously described (15) with the following modifications. KAPA HiFi HotStart ReadyMix (Roche) was used to amplify the DNA following ligations using the following conditions: 98°C for 45 seconds, followed by 8 cycles of 98°C for 15 seconds, 60°C for 30 seconds and 72°C for 30 seconds. The amplified DNA was purified using 1·8 X SPRI beads and eluted in 100 uL EB buffer (Qiagen). Ten uL of eluted DNA was used per well for targeted enrichment using multiplex, anchored hemi-nested PCR. Targeted sequencing was done as previously described on an Illumina NovaSeq 6000 instrument or a Complete Genomics T7 instrument (16).

### Mutation Analysis

To identify somatic mutations, a driver gene panel consisting of 121 commonly mutated regions in a total of 40 driver genes was evaluated (13,15). Only mutations present in both the Watson and Crick strands, absent in the matched normal, and defined as oncogenic were considered further. Mutations were defined as oncogenic if they were hotspots in oncogenes, or inactivating mutations (those predicted to truncate the protein product) in tumor suppressors genes -except for *TP53,* in which both inactivating mutations and hotspot mutations were defined as oncogenic (17–18). Hotspot mutations in *TP53* were defined as those which were non-synonymous and previously reported to be somatic in at least two different instances in the COSMIC database(19). Plasma DNA samples were scored as positive if there were two or more distinct mutant molecules identified in plasma DNA and there were no mutant molecules identified in the matched leukocyte DNA from the same patient. Positive scores in plasma could either reflect the presence of a tumor or some other clonal process, such as clonal hematopoiesis of indeterminate potential (CHIP) (13). To exclude CHIP with high confidence, we analyzed matched leukocytes from every patient for every mutation identified in the plasma sample. The number of leukocyte DNA molecules assessed was in excess of the number needed to ensure that the same mutation was *not* present with >99·99% probability based on a binomial distribution. In these distributions, the expected probability was set as the mutant allele fraction (MAF) observed in the plasma for that specific mutation. For example, if the MAF for a given mutation in the plasma was 0·02%, we evaluated >46047 leukocyte DNA molecules. The number of DNA molecules was calculated as the number of duplex UIDs observed in the sequencing, as there is a 1:1 correspondence between the number of duplex UIDs and the number of DNA molecules present in the library made from any DNA sample (15).

### Outcomes

The outcomes of interest were ctDNA positivity and a clinical cancer diagnosis. Only patients who had biopsy proven histopathologic diagnosis of invasion were classified as having cancer; patients with benign tumors, including non-invasive in situ tumors of the skin, were not classified as having cancer.

### Statistical analysis

Statistical tests were performed to assess whether there were differences in demographic, clinical and autoantibody characteristics between ctDNA-positive and ctDNA-negative patients. Detailed definitions and autoantibody assay methods are provided in the Supplement. The Student’s t-test was used for continuous variables and Fisher’s exact test for categorical variables. The Benjamini-Hochberg method was used to correct for multiple comparisons. All analyses were performed using R version 4·3·2.

## RESULTS

### Demographic and Disease Characteristics

A total of 134 patients were enrolled in the study, comprising 69 with SSc and 65 with DM. Both cohorts consisted predominantly of white females (64% female, 83% white for SSc; 78% female, 83% white for DM). The median age of rheumatic disease onset was 57 years (IQR 50-67) for SSc and 53 years (IQR 37-62) for DM [Tables 1-2]. These demographic characteristics are consistent with those of our overall cohorts. Among the 69 SSc patients, the most prevalent autoantibody was anti-POLR3 (n=28, 49%), followed by anti-RNPC3 (n=17, 30%), Ro52 (n=11, 19%), and anti-centromere, anti-Scl-70, and anti-Th/To (all n=8, 14%). In the 65 DM patients, anti-TIF1γ was the most frequent autoantibody (n=27, 43%), followed by anti-Ro52 (n=17, 26%) and anti-NXP2 (n=13, 21%) [Tables 1-2]. The median time from rheumatic disease symptom onset to the baseline ctDNA test was 1·3 years (IQR 0·8-1·9) for SSc and 1·4 years (IQR 1·0-2·6) for DM. The average follow-up time after the baseline ctDNA draw was 24 months for SSc and 18 months for DM.

**Table 1.**
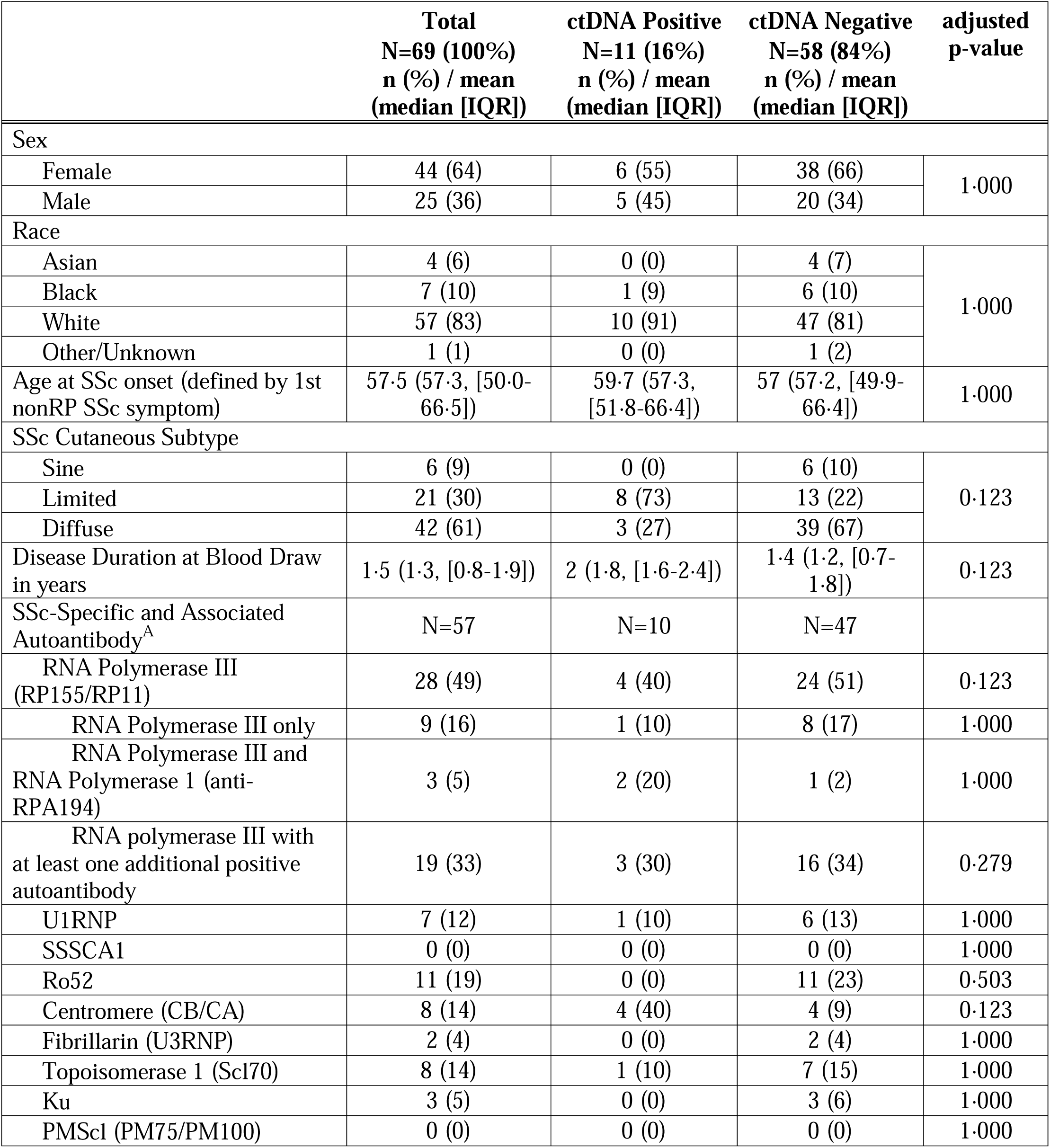
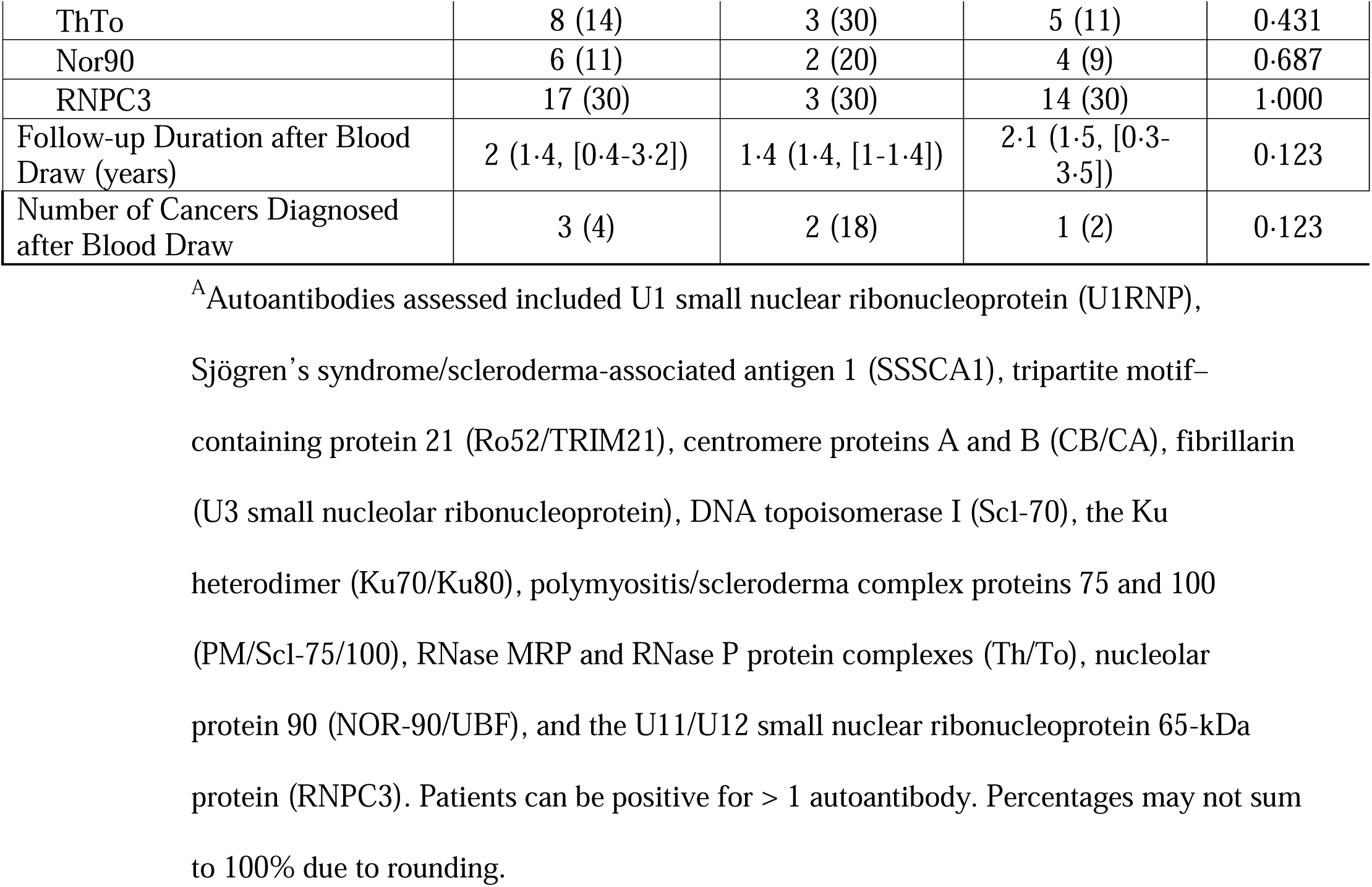
Systemic Sclerosis Cohort stratified by ctDNA Positivity at Baseline.

**Table 2.** Dermatomyositis Cohort Stratified by ctDNA Positivity at Baseline.

|  | **Total<br>N=65 (100%)<br>n (%) / mean<br>(median [IQR])** | **ctDNA Positive<br>N=6 (9%)<br>n (%) / mean<br>(median [IQR])** | **ctDNA Negative<br>N=59 (91%)<br>n (%) / mean<br>(median [IQR])** | **adjusted<br>p-value** |
|---|---|---|---|---|
| Sex |  |  |  |  |
| Female | 51 (78) | 6 (100) | 45 (76) | 1·000 |
| Male | 14 (22) | 0 (0) | 14 (24) |  |
| Race |  |  |  |  |
| Asian | 3 (5) | 1 (17) | 2 (3) | 1·000 |
| Black | 6 (9) | 1 (17) | 5 (8) |  |
| White | 53 (83) | 4 (67) | 49 (83) |  |
| Other/Unknown | 3 (5) | 0 (0) | 3 (5) |  |
| Age at IIM onset (Median, IQR) | 50·4 (53, [37·4-61·9]) | 62·7 (61·1, [59·5-69·1]) | 49·1 (52·1, [36·8-61·5]) | 0·230 |
| Disease Duration at Blood Draw | 2 (1·4, [1-2·6]) | 1·4 (0·9, [0·5-1·1]) | 2·1 (1·7, [1-2·6]) | 1·000 |
| Myositis-Specific and Associated Autoantibody<sup>A</sup> | N=63 | N=6 | N= 57 |  |
| TIF1g | 27 (43) | 2 (33) | 25 (44) | 1·000 |
| TIF1g+, CCAR1- and Sp4- | 6 (10) | NA (NA) | 6 (11) | 1·000 |
| TIF1g+, CCAR1+ or Sp4+ | 21 (33) | 2 (33) | 19 (33) | 1·000 |
| TIF1g+, CCAR1+ and Sp4+ | 9 (14) | 1 (17) | 8 (14) | 1·000 |
| NXP2 | 13 (21) | 2 (33) | 11 (21) | 1·000 |
| SAE | 2 (3) | 0 (0) | 2 (4) | 1·000 |
| Mi2 | 0 (0) | 0 (0) | 0 (0) | 1·000 |
| MDA5 | 6 (10) | 2 (33) | 4 (7) | 1·000 |
| Ro52 | 17 (26) | 1 (17) | 16 (31) | 1·000 |
| Jo-1 | 2 (3) | 0 (0) | 2 (4) | 1·000 |
| PL-7 | 0 (0) | 0 (0) | 0 (0) | 1·000 |
| PL-12 | 0 (0) | 0 (0) | 0 (0) | 1·000 |
| OJ | 0 (0) | 0 (0) | 0 (0) | 1·000 |
| EJ | 0 (0) | 0 (0) | 0 (0) | 1·000 |
| PM-Scl | 4 (6) | 0 (0) | 4 (7) | 1·000 |
| Ku | 2 (3) | 0 (0) | 2 (4) | 1·000 |
| SRP | 0 (0) | 0 (0) | 0 (0) | 1·000 |
| Follow-up Duration after Blood Draw | 1·5 (1·5, [0·7-2·4]) | 2·3 (2·4, [2·3-2·4]) | 1·4 (1·3, [0·6-2·2]) | 0·230 |
| Cancer Diagnosed after Blood Draw | 3 (5) | 2 (33) | 1 (2) | 0·284 |

<sup>A</sup>Autoantibodies assessed included transcription intermediary factor 1-gamma (TIF1γ), with subgroups defined by co-reactivity to cell cycle and apoptosis regulator protein 1 (CCAR1) and specificity protein 4 (Sp4) [TIF1γ□/CCAR1□/Sp4□; TIF1γ□/CCAR1□ or Sp4□; TIF1γ□/CCAR1□ and Sp4□], and by Ro52 (tripartite motif–containing protein 21) status (Ro52□ or Ro52□); nuclear matrix protein 2 (NXP2), small ubiquitin-like modifier activating enzyme (SAE), Mi-2 chromatin-remodeling complex, melanoma differentiation–associated protein 5 (MDA5), tripartite motif–containing protein 21 (Ro52), histidyl-tRNA synthetase (Jo-1), threonyl-tRNA synthetase (PL-7), alanyl-tRNA synthetase (PL-12), isoleucyl-tRNA synthetase (OJ), glycyl-tRNA synthetase (EJ), polymyositis/scleroderma complex (PM/Scl), Ku heterodimer (Ku70/Ku80), and signal recognition particle (SRP). Patients can be positive for > 1 autoantibody. Percentages may not sum to 100% due to rounding.

### Somatic mutations

After excluding 68 mutations attributable to clonal hematopoiesis of indeterminate potential (CHIP; 45 mutations in 23 patients in the SSc cohort, 23 mutations in 9 patients in the DM cohort), 16% (11/69) of SSc patients and 9% (6/65) of DM patients had at least one mutation that was scored positive at baseline. There were no significant differences in sex, race, or ethnicity between ctDNA-positive and negative patients [Tables 1-2]. After correction for multiple comparisons, no non-neoplastic disease feature or disease activity measure differed significantly between ctDNA-positive and ctDNA-negative patients [Tables 1-2, Supplemental Tables 1-2]. However, there were several interesting trends including ctDNA-positive patients with SSc were more likely to have limited SSc (73% vs 22%, p=0·123), a lower mean modified Rodnan skin score (mRSS) at the time of ctDNA blood draw (6 vs 11, p=0·322), and positive anti-centromere autoantibodies (40% vs 9%, p=0·123) [Table 1, Supplemental Table 1]. In both SSc and DM, there were no differences in immunosuppressive treatment regimens by ctDNA positivity [Supplemental Tables 3-4].

Among both diseases, mutations in the *TP53* gene were the most prevalent at baseline. In SSc, eight out of 11 (73%) patients who were ctDNA positive had a *TP53* mutation, and one patient each (9%) had mutations in *PTEN*, *PIK3CA* and *FGFR3*. In the DM cohort, three of six patients (50%) harbored *TP53* mutations, and one patient each (17%) had mutations in *PTEN*, *PIK3CA* and *EGFR*. Among the 11 SSc ctDNA-positive patients, two were diagnosed with cancer within a median follow-up of 1·4 years [Tables 3-4]. Among the 6 DM ctDNA-positive patients on baseline testing, one was diagnosed with cancer within a median follow-up of 2·4 years. In approximately half of the patients (n=27 in SSc and n=46 in DM), a second blood draw was obtained and available for analysis. Among the 17 patients who were ctDNA-positive at baseline, 11 (five SSc, six DM) had a second sample available. In SSc, none of the five (0%) patients had a positive second sample (and none had cancer), whereas in DM, two of six (33%) patients remained positive for the same mutation(s). Of the two DM patients who were positive in both the first and second samples, one later developed cancer (MYOS 59), while the other patient remained cancer-free after 2·5 years of follow-up. Additional data on somatic mutations on the subgroup of patients with a second ctDNA measurement are found in the Supplement. Figure 1 illustrates the timeline of ctDNA measurements relative to one another, the follow-up period, and cancer outcomes. Details regarding cancer screening tests performed on both cohorts are recorded in Supplemental Tables 5-6.

**Fig 1.**
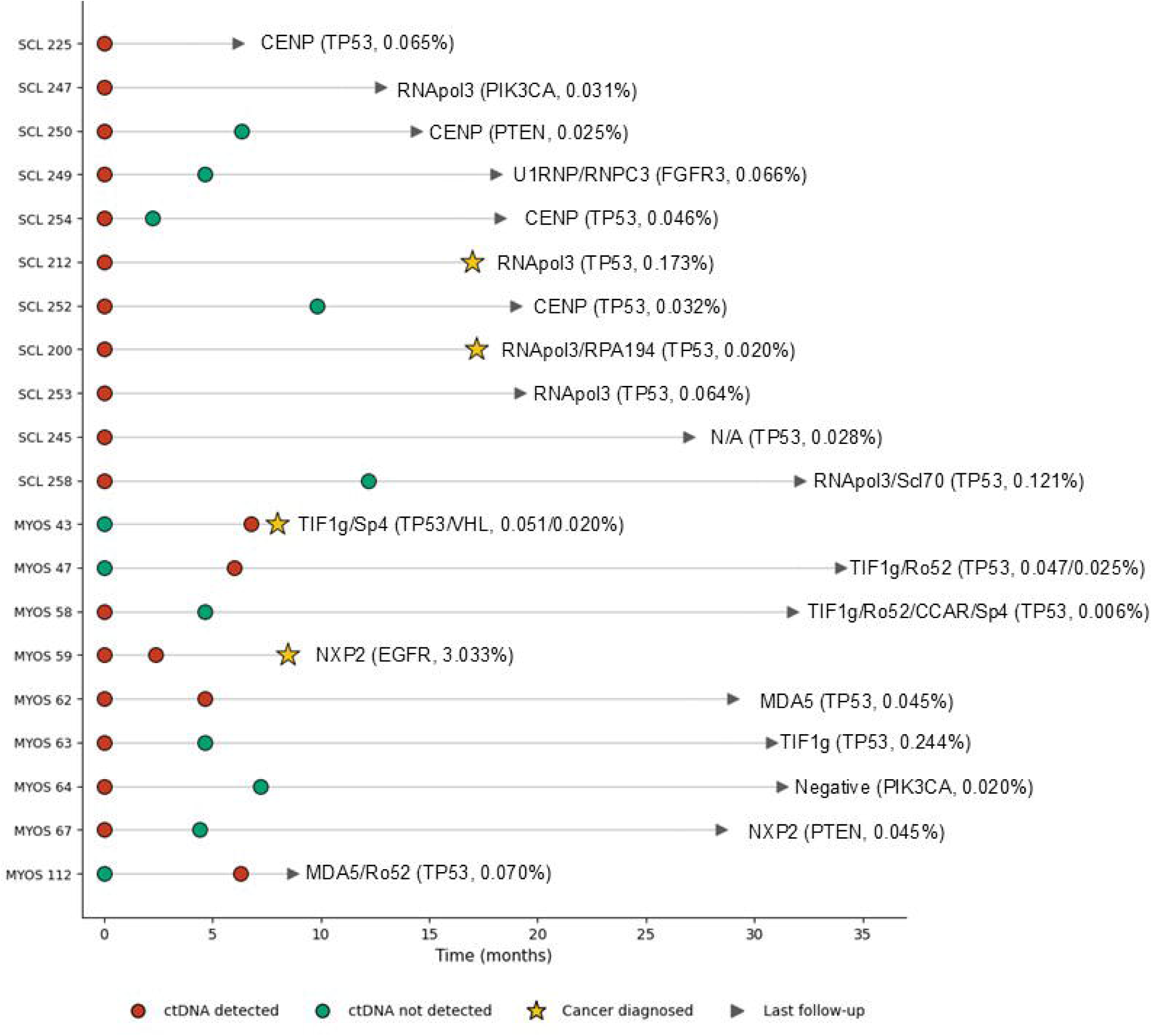
Follow-up duration and cancer outcome in 20 SSc/DM patients with mutations. Of the 17 patients with mutations detected at baseline, 11 had repeat ctDNA measurements. Of these 11, 2 remained positive and 9 converted to negative. Three patients who were initially negative tested positive at the second time point, one of whom went on to be diagnosed with cancer (MYOS43). The autoantibody is listed for each patient, followed by the mutated gene(s) and mutant allele frequency in parentheses.

**Table 3.**
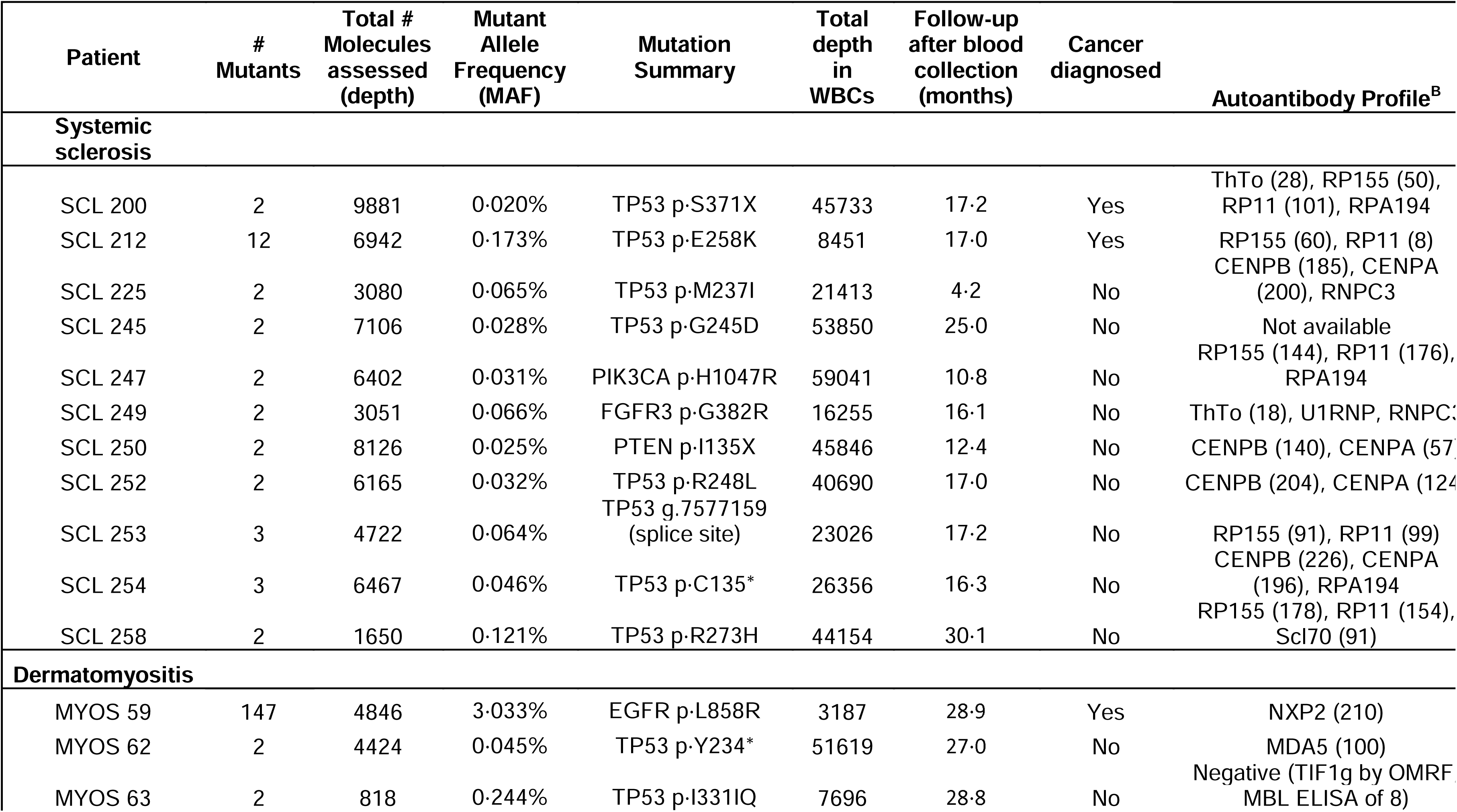
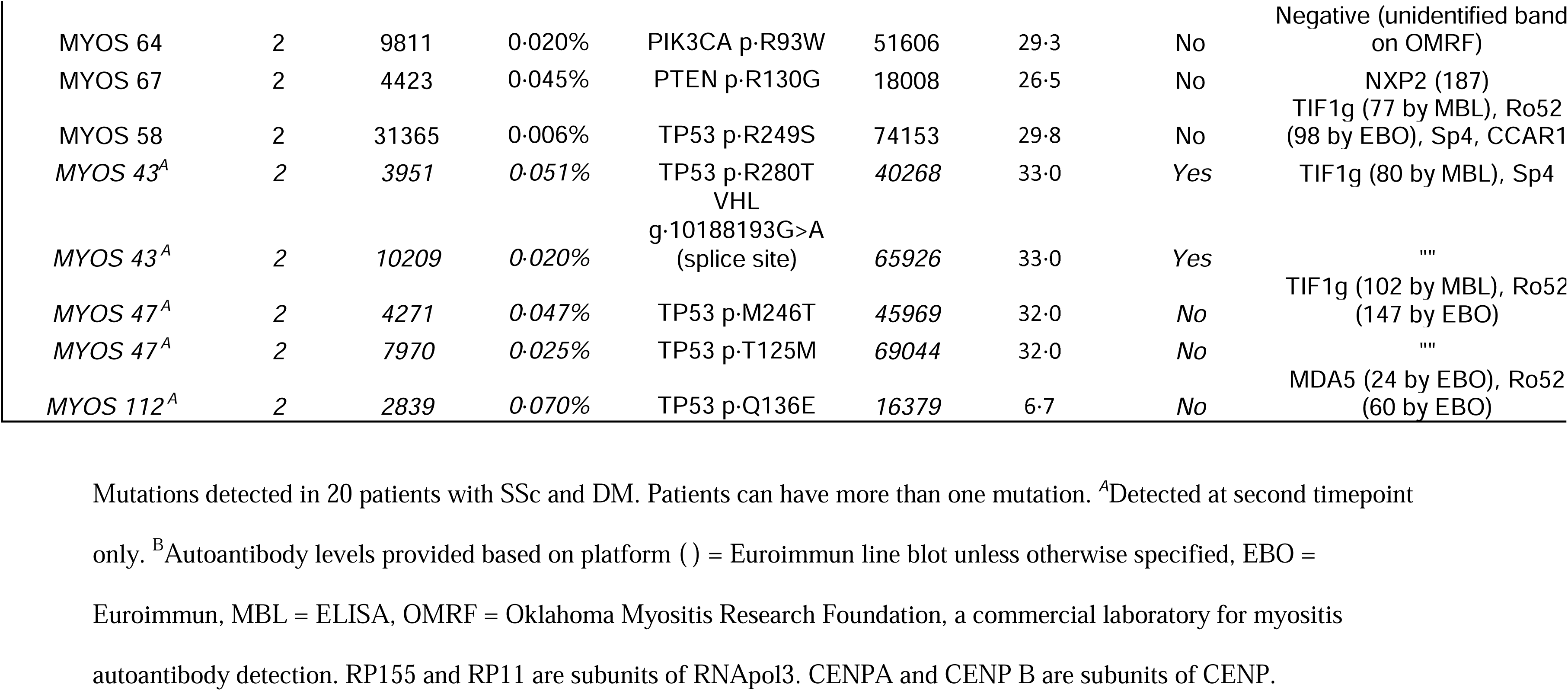
Mutations detected in ctDNA.

**Table 4.** Clinically diagnosed cancers by site, type, ctDNA result, and autoantibody status.

| **Rheumatic Disease** | **Age Window at Rheumatic Disease Onset/Sex** | **Cancer Risk Factors (smoking, EtOH)** | **Cancer Site** | **Histology** | **Time from Blood Draw to Cancer Diagnosis (months)** | **AJCC Stage** | **Positive by ctDNA (mutated gene, MAF)** | **Autoantibody<sup>A</sup>** | **Matched Mutation present in Tumor** |
|---|---|---|---|---|---|---|---|---|---|
| Systemic sclerosis |  |  |  |  |  |  |  |  |  |
| SCL 200 | 70-75/M | smoking (quit 2016, 54 pack years), social EtOH | External ear | Squamous cell carcinoma with perineural invasion | 17·0 | NR | Yes, TP53 (0·020%) | RNApol3, RPA194 | No |
| SCL 208 | 50-55/M | 12-16 EtOH/wk | Prostate | Adenocarcinoma | 64·6 | II/Gleason 7 | No | Th/To, RNApol3 | N/A |
| SCL 212 | 56-60/M | smoking (1 pack/day), 6 EtOH/wk | Lung and Bronchus | Squamous cell carcinoma | 17·2 | IV | Yes; TP53 (0·173%) | [RNApol3] | Yes |
| Dermatomyositis |  |  |  |  |  |  |  |  |  |
| MYOS 59 | 70-75/F | smoking (quit 2001, 15 pack years), social EtOH | Lung and Bronchus | Adenocarcinoma, NOS | 8·5 | III | Yes, EGFR (3·03%), +aneuploidy | NXP2, Sp4 | Yes |
| MYOS 107 | 60-65/M | 3 EtOH/wk | Tonsil | Papillary squamous cell carcinoma | 4·0 | I | No | TIF1g | N/A |
| MYOS 43 | 76-80/M | social EtOH | Urinary Bladder | Carcinoma, NOS | 8·0/1·4 | Ic | No on 1st draw, Yes on 2nd draw TP53 (0·051%) | TIF1g, Sp4 | Yes |

<sup>A</sup>Brackets [ ] indicate clinically positive results that do not meet Euroimmun line blot criteria as described in the Methods. MAF = mutant allele frequency; EtOH = alcohol use; NOS = not otherwise specified. TP53 and EGFR denote the genes assessed.

### Cancer Detection

In the SSc cohort, three of 69 patients (4·3%) were diagnosed with cancer during follow-up, including a prostate cancer (Gleason grade 7), a stage IV lung squamous cell carcinoma, and an invasive squamous cell carcinoma of the ear with perineural invasion [Table 4]. Two of the three patients with cancer had detectable ctDNA: One patient, SCL 212, had a detectable mutation (*TP53* p·E258K with mutant allele frequency (MAF) 0·173%; Table 3) 17 months prior to a Stage IV lung cancer diagnosis. Interestingly, this patient had a negative PET/CT 16 months prior to lung cancer diagnosis despite ctDNA positivity. Analysis of the corresponding tumor tissue from this patient identified the same mutation that was detected in plasma. The second patient, SCL 200, had a positive mutation (*TP53* p·S371X with MAF 0·02%; Table 4) 17 months prior to being diagnosed with an invasive squamous cell carcinoma of the ear; this mutation was not detected in the corresponding cancer tissue. We did not detect ctDNA in the patient with prostate cancer, SCL 208, in a plasma sample taken 65 months prior to diagnosis.

In the DM cohort, three of 65 patients (4·6%) were diagnosed with cancer, which included a stage III lung adenocarcinoma, a stage I squamous cell carcinoma (SCC) of the tonsils, and a stage I bladder carcinoma [Table 4]. The patient with lung adenocarcinoma, MYOS 59, had a positive ctDNA test (*EGFR* p·L858R with MAF at 3·03%; Table 3) 8·5 months prior to lung cancer diagnosis. The *EGFR* mutation was identified in the corresponding primary tumor. This patient underwent a contrast-enhanced chest CT seven months before diagnosis that identified a pulmonary nodule with a clinical recommendation for repeat imaging in one year. The patient who was diagnosed with bladder cancer, MYOS 43, had negative ctDNA eight months prior to cancer diagnosis, but a positive ctDNA test one month prior to cancer diagnosis. Two mutations were identified at this timepoint: a *TP53* p·R280T mutation, which was also found in the matched tumor, and a *VHL* splice site mutation that was not identified in the primary tumor. This patient had a negative PET/CT scan six months before diagnosis. In the patient with tonsillar SCC, MYOS107, ctDNA was not detectable in a blood sample collected four months prior to cancer diagnosis.

Across both diseases, four of six patients who developed cancer had a positive ctDNA test (two in SSc, two in DM), whereas the assay was positive in 16 of the 128 patients who remained cancer-free during follow-up (nine in SSc, seven in DM) [Figure 2]. Thus, ctDNA positivity was associated with a 11·4-fold higher risk of subclinical malignancy compared with ctDNA negativity, with a negative predictive value of 98% and a positive predictive value of 20%.

**Fig. 2.**
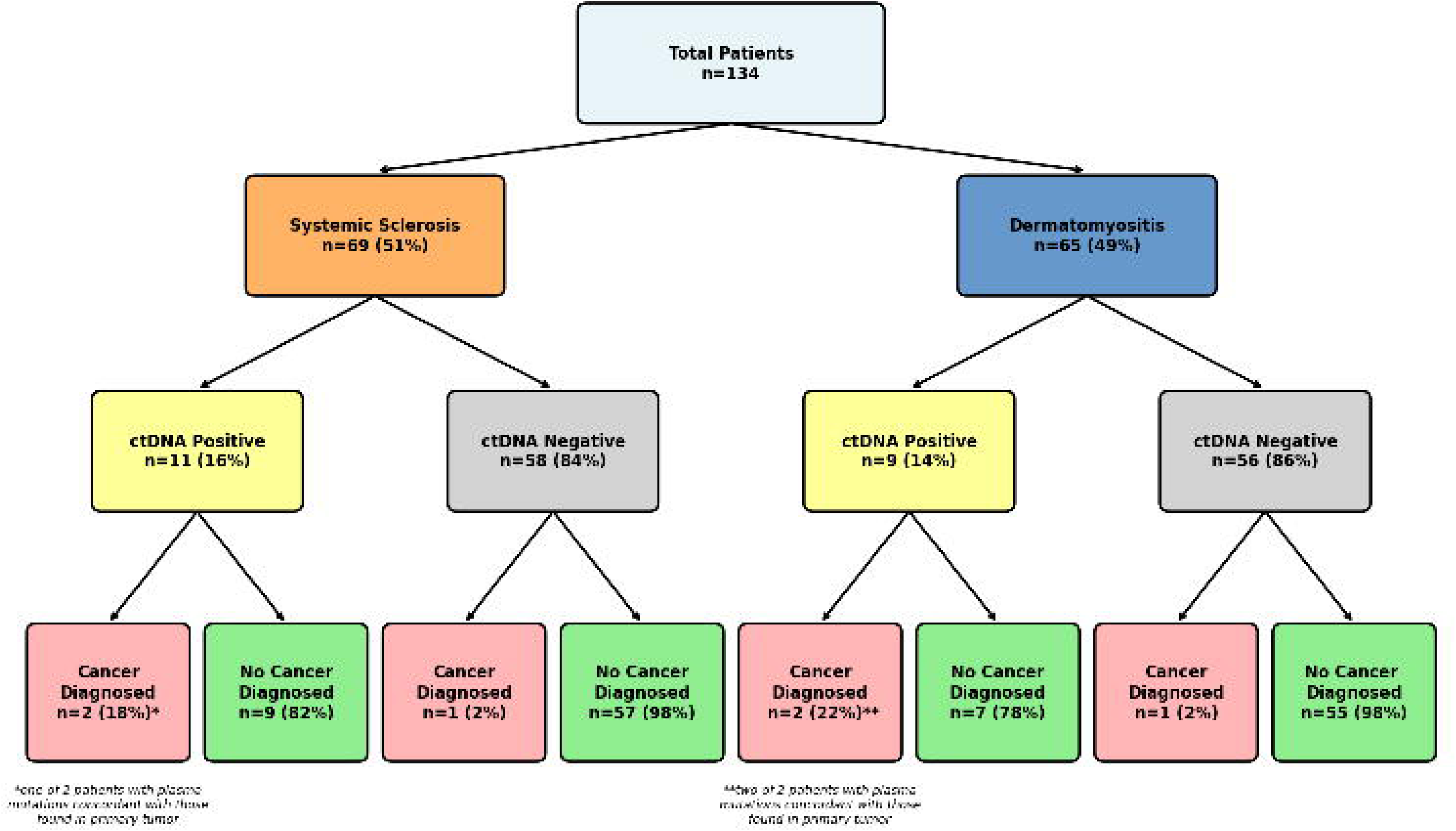
ctDNA positivity (ever/never) and cancer occurrence during follow-up in scleroderma and dermatomyositis cohorts. Among 134 patients (69 with systemic sclerosis [SSc] and 65 with dermatomyositis [DM]), 11 (16%) and 9 (14%) had positive ctDNA, respectively. During follow-up, cancer was diagnosed in 18% of ctDNA-positive SSc patients and 22% of ctDNA-positive DM patients, compared to 2% of ctDNA-negative patients. Overall, ctDNA positivity was associated with a 11.4-fold increased risk of occult malignancy compared with ctDNA negativity and showed a negative predictive value of 98% and a positive predictive value of 20% for cancer within the observation window.

## DISCUSSION

Compelling epidemiologic and mechanistic studies suggest that cancer may drive the development of autoimmunity in subgroups of patients with SSc and DM identified by distinct autoantibody responses. However, most patients within these autoantibody subgroups do not have a clinically diagnosed cancer. In this study, we detected evidence of cancer, through analysis of ctDNA in blood, in four of six patients who developed a clinically detectable cancer at a median of 13 months prior to clinical diagnosis. Of note, in two of these 4 cancers, diagnostic PET/CTs around the time of ctDNA positivity were unremarkable, highlighting the potential value of ctDNA assessment as a cancer screening tool.

One striking finding of our work is that of 20 patients with detectable ctDNA mutations, only four developed a clinically detectable cancer despite intensive cancer surveillance over the ensuing months. And in one (SCL 200) of the four patients with a positive ctDNA test, the mutation was *not* identified in the patient’s subsequent cancer biopsy tissue. Though the nominal positive predictive value (PPV 20%) of our test was higher than that for mammography for breast cancer or spiral CT for lung cancer, it is still lower than optimal from an interventional standpoint. There are at least two hypotheses to explain the low PPV. The most intellectually exciting of these is that more of these ctDNA-positive patients actually had cancer, but the cancer was not large enough to cause symptoms or to be detected by routine surveillance measures. According to this hypothesis, some of the ctDNA-positive patients will be diagnosed with cancer in the future. Indeed, ctDNA can be detected up to three years prior to clinical cancer diagnosis (13), and the median follow-up time for patients in the current study was only 17 months. The presence of cell-free DNA but the absence of clinically detectable cancer could accordingly represent a state of equilibrium between the immune system and cancer (7). This hypothesis is consistent with the underlying rationale for our study, i.e., the immune response elicited by the tumor is sufficient to inhibit its progression, sometimes to the extent that clinical cancer never appears.

The other hypotheses explain the ctDNA-positives as either technical or biological artifacts, so there is no cancer at all present in the patients with positive ctDNA results. When the same ctDNA test is applied to young healthy individuals, false positives are rare, so technical artifacts are minimized (15). However, biological “artifacts” are still possible. The most likely type of biological artifact is clonal hematopoiesis of indeterminate potential (CHIP). Though we evaluated very large numbers of molecules to confidently exclude leukocyte DNA mutations in even a minor fraction of leukocytes, we could not exclude mutations that arose from an even smaller fraction of lymphocytes, such as those that contributed cfDNA in excess of their representation in peripheral blood counts. Prior studies using DNA methylation have indeed shown that B-lymphocytes contribute more cfDNA than expected from their peripheral blood cell fraction (20). Moreover, other studies have suggested a higher prevalence of clonal hematopoiesis of CHIP in patients with autoimmune disease (21–22). Longitudinal monitoring of circulating tumor DNA mutations, paired with clinical phenotyping of patients’ disease activity and autoantibody profiles, is currently underway to test these relationships. A noteworthy strength of our study design was to purposefully focus on patients with newly diagnosed autoimmune disease (<3 years), as this is the population at highest risk of cancer diagnosis. Additionally, our inclusion of patients with a broad range of autoantibodies with different epidemiologic cancer associations was informative. Multiple epidemiologic studies have shown that specific autoantibody profiles stratify cancer risk within the first three years after rheumatic disease onset, with some profiles associated with increased risk, others with decreased risk, and others with no change (1, 2). However, several of the ctDNA positive patients had autoantibodies, such as anti-centromere, that have been linked to a decreased cancer risk in cohort studies or when compared to the general population. A potential explanation is that these autoantibody specificities are a surrogate for a highly effective anti-tumor immune response, accounting for the negative association observed in prior epidemiologic studies. It is noteworthy that among the six SSc patients who had an initial positive ctDNA test followed by a negative test, three had anti-centromere antibodies (SCL250, SCL252, SCL254).

Our study has several limitations. While we tried to employ systematic cancer screening strategies, not all patients underwent recommended tests in part due to the Covid-19 pandemic, and in part due to patient preference. Only a subset of patients had a second sample available for ctDNA analysis, and an important priority is examining the dynamic relationship between ctDNA levels, evolution of the immune response, and cancer emergence over time. As noted above, the follow-up time is relatively short, so the false-positive rate may be overestimated. And the relatively small number (n=six) of patients that developed cancer in this cohort engender wide confidence limits in all of the reported outcomes.

What are the potential clinical implications of our findings? From a rheumatology perspective, the earlier detection and treatment of cancer may lead to improvement, if not remission, of the rheumatic disease 23-24). From an oncologic perspective, the earlier detection of cancer could lead to improved survival, and even cure, of the cancers. There is evidence that a delay of even a month between diagnosis of a cancer and its surgical removal has a substantial effect on survival (25–27). And there is also much data indicating that anti-cancer drugs of all types are much more likely to induce prolonged remissions or cures when a smaller tumor burden is present at the time of treatment (28–30). Conversely, the relatively high ctDNA positive rate observed in our study is concerning, as it could lead to additional anxiety, or needless procedures, in patients who already have a serious autoimmune disease.

In summary, we have unequivocally shown that the cancers in some patients with specific types of autoimmune diseases can be detected with a blood test before conventional methods are able to detect them. This study thereby sets the stage for larger, prospective studies to determine whether such testing can benefit patients.

## Data Availability

Data and code used in the analysis will be made available in controlled-access repositories. A subset of participants did not permit sharing of their genetic data; therefore, the number of participants represented in the repositories will not exactly match the number included in publication. Accession numbers for data will be provided upon journal acceptance of the manuscript.

## Contributors

Conceptualization: CAM, YW, AR, BV, AAS. Investigation: CAM, YW, JSK, XW, AW, CD SDC, MP, JP, LD, NS, MBS, LF, GL, LKH, LCS, JJP, JA, FMW, AR, LCR, CB, NP, KWK, BV, AAS. Funding acquisition: CAM, KWK, BV, AAS. Supervision: BV, AAS. Writing – original draft: CAM, YW, BV, AAS. Writing – review & editing: CAM, YW, JSK, XW, AW, CD, SDC, MP, JP, LD, NS, MBS, LF, GL, LKH, LCS, JJP, JA, FMW, AR, LCR, CB, NP, KWK, BV, AAS.

## Data Sharing

Data and code used in the analysis will be made available in controlled-access repositories. A subset of participants did not permit sharing of their genetic data; therefore, the number of participants represented in the repositories will not exactly match the number included in publication. Accession numbers for data will be provided upon acceptance of manuscript.

## Acknowledgements

We thank Deborah Cummings-Thomas for her technical assistance with this project, and Laura Gutierrez, Qingyuan Yang and David Hines for performing antibody assays in the P30-funded Rheumatic Diseases Research Core Center.

## Funding

National Institutes of Health grant R01-AR073208 (AAS, LCR)

National Institutes of Health grant R01-AR083912 (CM)

National Institutes of Health grant K24-AR080217 (AAS)

National Institutes of Health grant P30-AR070254 (AR)

The Scleroderma Research Foundation

The Donald B. and Dorothy L. Stabler Foundation

The Chresanthe Staurulakis Memorial Fund

The Sara and Alex Othon Research Fund

The Nancy and Joachim Bechtle Precision Medicine Fund for Scleroderma

The Hyuai and Siuling Zhang Discovery Fund Dr. Peter Buck Fund

The Jerome L. Greene Foundation

The Lustgarten Foundation for Pancreatic Cancer Research

The Sol Goldman Sequencing Facility at Johns Hopkins

The Virginia and D.K. Ludwig Fund for Cancer Research

## AI Disclosure

AI software (Claude 5 Sonnet, Anthropic) was used to proofread the manuscript for grammar and clarity after its completion, and to assist in writing code used to generate draft plots for Figures 1 and 2. All AI-assisted outputs were reviewed and verified by the authors, who take full responsibility for the content of the manuscript.

