## Supplement for "Circulating Tumor DNA in Patients with Early-Onset Systemic Sclerosis and Dermatomyositis"

### SUPPLEMENTAL METHODS

#### Autoantibodies

Anti-Mi2 was considered positive if both Mi2a and Mi2b were ≥15/+; if either Mi2a or Mi2b was positive, sera were tested for anti-Mi2 by ELISA (MBL, Nagoya, Japan) to verify the result. Similarly, anti-PM-Scl was considered positive if PM75 and PM100 were both ≥15/+. Given concerns of suboptimal sensitivity and specificity for anti-TIF1γ and anti-NXP2 as assayed by Euroimmun (48), these autoantibodies were systematically also assayed by ELISA (for anti-TIF1γ; MBL, Nagoya, Japan) and immunoprecipitation (for anti-NXP2, using $^{35}$S-methionine-labeled NXP2 generated by in vitro transcription and translation as input).

#### Disease activity and damage indices

Disease activity and damage in systemic sclerosis (SSc) were assessed using validated clinical indices and objective measures reflecting cutaneous, musculoskeletal, and cardiopulmonary involvement (49). Overall disease severity was quantified using a modified Medsger Disease Severity Scale, which evaluates organ systems including general, peripheral vascular, skin, muscle, gastrointestinal, and lung involvement. Each domain is scored from 0, indicating no involvement, to 4, indicating end-stage disease, with higher scores reflecting greater organ-specific disease severity. Skin involvement was assessed using the modified Rodnan skin score (mRSS), which grades skin thickness at 17 body sites on a scale from 0 to 3, yielding a total score ranging from 0 to 51. Higher mRSS values indicate more extensive and severe cutaneous involvement and are commonly used as a marker of disease activity, particularly in early or diffuse SSc. Musculoskeletal involvement was further characterized by the presence or absence of tendon friction rubs, assessed by physical examination at standard anatomical sites, which were considered a marker of active disease and more severe disease phenotype. Pulmonary involvement was evaluated using pulmonary function testing, including forced vital capacity (FVC) and diffusing capacity for carbon monoxide (DLCO), expressed as percent predicted based on age, sex, height, and race. Interstitial lung disease was defined by the presence of radiographic fibrosis on high-resolution chest CT, chest radiograph, or rales on physical examination.

Disease activity and damage in dermatomyositis (DM) were assessed using validated clinician-reported, patient-reported, laboratory, and functional outcome measures encompassing both cutaneous and muscular disease domains (50). Cutaneous disease activity and damage were measured using the Cutaneous Dermatomyositis Disease Area and Severity Index (CDASI), in which activity scores capture the extent and severity of inflammatory skin features such as erythema, scale, excoriation, and ulceration across multiple anatomical sites, while damage scores reflect chronic and irreversible cutaneous changes, including poikiloderma, calcinosis, and scarring. Overall disease activity was additionally assessed using physician and patient global assessments, each recorded on a scale ranging from 0, indicating no disease activity, to 10, indicating the most severe disease activity imaginable. Muscular involvement was evaluated using the Manual Muscle Testing-8 (MMT-8), which assesses strength in eight bilateral proximal muscle groups, with total scores ranging from 0 to 150 and lower scores indicating greater muscle weakness and higher disease activity. Serum creatine kinase (CK) levels were measured as a biochemical marker of muscle inflammation and injury. Patients were typically seen at 4- to 6-month intervals, consistent with the standard follow-up schedule at our centers.

**Supplemental Table 1. Systemic Sclerosis Disease Activity Stratified by ctDNA Positivity at Baseline**

| SSc Disease Activity measures | Total<br>N=69 (100%)<br>n (%) / mean (median [IQR]) | ctDNA Positive<br>N=11 (16%)<br>n (%) / mean (median [IQR]) | ctDNA Negative<br>N=58 (84%)<br>n (%) / mean (median [IQR]) | adjusted p-value |
|---|---|---|---|---|
| Modified Rodnan Skin Score (mRSS) | 10.2 (8, [3-17]) | 6.1 (4, [3-8.5]) | 11 (8.5, [3-17.8]) | 0.322 |
| Forced vital capacity percent predicted (ppFVC) | 93.4 (95.2, [79.2-109.9]), N=53 | 86.3 (91.4, [73.7-95.3]), N=8 | 94.6 (95.5, [80.6-111.7]), N=45 | 1.000 |
| ppFVC < 70% | 9 (17), N=53 | 2 (25), N=8 | 7 (16), N=45 | 1.000 |
| Diffusing capacity percent predicted (ppDLCO) | 78.1 (79, [61.9-94.5]), N=48 | 92.3 (82.4, [71.2-119.7]), N=7 | 75.7 (76, [62-94.3]), N=41 | 1.000 |
| ppDLCO < 70% | 16 (33), N=48 | 2 (29), N=7 | 14 (34), N=41 | 1.000 |
| C-reactive protein (CRP) mg/L | N=31 | N=8 | N=23 | 1.000 |
| <0.3 | 8 (26) | 3 (38) | 5 (22) | |
| 0.3-<1.0 | 13 (42) | 1 (12) | 12 (52) | |
| 1.0-10.0 | 10 (32) | 4 (50) | 6 (26) | |
| >10.0 | 0 (0) | 0 (0) | 0 (0) | |
| Tendon friction rubs | 14 (23), N=61 | 2 (22), N=9 | 12 (23), N=52 | 1.000 |
| Moderate/Severe RP Medsger Severity Score at Baseline[A] | 7 (10) | 0 (0) | 7 (12) | 1.000 |
| Moderate/Severe General Medsger Severity Score at Baseline | 17 (27), N=63 | 2 (20), N=10 | 15 (28), N=53 | 1.000 |
| Moderate/Severe GI Medsger Severity Score at Baseline | 6 (9) | 1 (9) | 5 (9) | 1.000 |
| Moderate/Severe Kidney Medsger Severity Score at Baseline | 7 (10), N=67 | 0 (0) | 7 (12), N=56 | 1.000 |
| Moderate/Severe Muscle Medsger Severity Score at Baseline | 4 (6), N=65 | 1 (11), N=9 | 3 (5), N=56 | 1.000 |
| Moderate/Severe RP Medsger Severity Score (maximum) | 11 (16) | 1 (9) | 10 (17) | 1.000 |
| Moderate/Severe General Medsger Severity Score (maximum) | 20 (30), N=66 | 2 (18) | 18 (33), N=55 | 1.000 |
| Moderate/Severe GI Medsger Severity Score (maximum) | 33 (48) | 5 (45) | 28 (48) | 1.000 |

| | | | | |
|---|---|---|---|---|
| Moderate/Severe Kidney Medsger Severity Score (maximum) | 8 (12) | 0 (0) | 8 (14) | 1.000 |
| Moderate/Severe Muscle Medsger Severity Score (maximum) | 7 (10), N=68 | 1 (10), N=10 | 6 (10) | 1.000 |
| Maximum mRSS over follow-up | 13.2 (11, [4-19]) | 9.1 (4, [3-13]) | 14 (13.5, [4-20.5]) | 1.000 |
| ILD | 23 (33) | 3 (27) | 20 (34) | 1.000 |
| Telangiectasia | 45 (65) | 8 (73) | 37 (64) | 1.000 |
| Calcinosis | 5 (7), N=67 | 0 (0), N=10 | 5 (9), N=57 | 1.000 |
| Myopathy | 12 (18), N=67 | 1 (10), N=10 | 11 (19), N=57 | 1.000 |
| Renal crisis | 10 (14) | 0 (0) | 10 (17) | 1.000 |

[A]Medsger Severity Scores were dichotomized, with moderate/severe disease equating to scores 2, 3, or 4.

ILD = radiographic changes on CT chest, radiograph, or rales on physical examination.

**Supplemental Table 2**. **Dermatomyositis Disease Activity Stratified by ctDNA Positivity at Baseline**

| DM Disease Activity | Total N=65 (100%) n (%) / mean (median [IQR]) | ctDNA Positive N=6 (9%) n (%) / mean (median [IQR]) | ctDNA Negative N=59 (91%) n (%) / mean (median [IQR]) | adjusted p-value |
|---|---|---|---|---|
| CDASI activity | 11.8 (10, [3-16.5]), N=64 | 9.3 (6, [3.5-13]) | 12 (10, [3.2-17.5]), N=58 | 0.571 |
| CDASI damage | 1.6 (0, [0-2]), N=64 | 1 (0, [0-1.5]) | 1.7 (0, [0-1.8]), N=58 | 0.571 |
| Physician Global | 3.7 (3, [2-5]), N=54 | 3 (3, [1.5-3.8]) | 3.8 (3, [2-5.2]), N=48 | 0.571 |
| Patient Global | 4.1 (4, [2-6]), N=58 | 3.5 (4, [2.5-4.8]) | 4.2 (4, [2-6]), N=52 | 0.571 |
| MMT8 | 147.3 (150, [148-150]), N=59 | 147.8 (149, [148-150]) | 147.2 (150, [148-150]), N=53 | 0.729 |
| CK (+/- 6 weeks) | 175.7 (82, [49.5-106.5]), N=55 | 475.7 (82.5, [42.5-158.5]) | 138.9 (82, [52-105]), N=49 | 0.571 |

CDASI = cutaneous dermatomyositis disease area and severity index; MMT8 = Manual muscle testing-8; CK = creatine kinase.

**Supplemental Table 3**. **Immunosuppressive/Immunomodulatory medication use at the time of Baseline Blood Draw in Systemic Sclerosis Patients**

| **Medication** | **ctDNA Positive (n=11)**<br>**n (%)** | **ctDNA Negative (n=58)**<br>**n (%)** | **adjusted p-value** |
|---|---|---|---|
| Mycophenolate mofetil | 5 (45%) | 36 (62%) | 0.692 |
| Tocilizumab | 0 (0%) | 2 (3%) | 1.000 |
| Methotrexate | 1 (9%) | 6 (10%) | 1.000 |
| Intravenous immunoglobulin | 1 (9%) | 1 (2%) | 0.692 |
| Hydroxychloroquine | 1 (9%) | 10 (17%) | 1.000 |
| Prednisone | 0 (0%) | 10 (17%) | 0.692 |
| Rituximab | 0 (0%) | 1 (2%) | 1.000 |
| None | 5 (45%) | 13 (22%) | 0.692 |
| **Number of immunosuppressants** | | | |
| Range | 0-2 | 0-4 | |
| Mean (SD) # of immunosuppressants | 0.73 (0.79) | 1.14 (0.89) | |
| Median # of immunosuppressants | 1 | 1 | |

**Supplemental Table 4. Immunosuppressive/Immunomodulatory medication use at the time of Baseline Blood Draw in Dermatomyositis Patients**

| Medication | ctDNA Positive (n=6) n (%) | ctDNA Negative (n=59) n (%) | adjusted p-value |
|---|---|---|---|
| Corticosteroids | 5 (83%) | 25 (42%) | 0.479 |
| Methotrexate | 4 (67%) | 12 (20%) | 0.319 |
| Azathioprine | 0 (0%) | 2 (3%) | 1.000 |
| Mycophenolate | 0 (0%) | 18 (31%) | 0.642 |
| Tacrolimus | 1 (17%) | 4 (7%) | 1.000 |
| Intravenous immunoglobulin | 2 (33%) | 16 (27%) | 1.000 |
| Rituximab | 0 (0%) | 12 (20%) | 1.000 |
| Hydroxychloroquine | 0 (0%) | 12 (20%) | 1.000 |
| Tofacitinib | 0 (0%) | 1 (2%) | 1.000 |
| Unknown | 0 (0%) | 1 (2%) | 1.000 |
| None | 0 (0%) | 6 (10%) | 1.000 |
| **Number of immunosuppressants** | | | |
| Range | 2 | 4 | |
| Mean # of immunosuppressants | 2 | 1.79 | |
| Median # of immunosuppressants | 2 | 2 | |

**Supplemental Table 5. Cancer screening tests performed in systemic sclerosis cohort**

| USPSTF Recommended Screening At the Time of Blood Draw[A] | | |
|---|---|---|
| | Test Completion (ctDNA Positive) | Test Completion (ctDNA Negative) |
| Mammogram | 60% | 94% |
| Pap Smear | 50% | 79% |
| Chest CT | 100% | 100% |
| Colonoscopy | 88% | 98% |
| **Additional Cancer Screening Performed From SSc Onset Onwards[B]** | | |
| Chest CT | 100% | 93% |
| Abdomen/Pelvis CT | 55% | 62% |
| PET/CT | 27% | 47% |
| PSA | 80% | 60% |

[A]USPSTF Screenings: Each row shows the percentage of eligible patients who completed the specified cancer screening test (number completed divided by number eligible). Eligibility criteria follow USPSTF guidelines; for example, only women aged ≥40 years are included in the denominator for mammography. Colonoscopy was considered completed if performed within 2 years of sample collection, whereas all other USPSTF-recommended screenings were required to be completed within 6 months of sample collection.
[B]Additional Cancer Screenings: All additional screening tests were required to be performed within 6 months before or any time after systemic sclerosis (SSc) symptom onset.

**Supplemental Table 6.** Cancer screening tests performed in dermatomyositis cohort.

| **USPSTF Recommended Screening At the Time of Blood Draw**[A] | | |
|---|---|---|
| | Test Completion (ctDNA Positive) | Test Completion (ctDNA Negative) |
| Mammogram | 80% | 79% |
| Pap Smear | 100% | 71% |
| Chest CT per USPSTF | N/A | 50% |
| Colonoscopy | 60% | 90% |
| **Additional Cancer Screening Performed From DM Onset Onwards**[B] | | |
| Transvaginal Ultrasound | 33% | 42% |
| Chest CT | 100% | 97% |
| Abdomen/Pelvis CT | 100% | 76% |
| PET/CT | 50% | 10% |
| PSA | 100% | 64% |

[A]USPSTF Screenings: Each row shows the percentage of eligible patients who completed the specified cancer screening test (number completed divided by number eligible). Eligibility criteria follow USPSTF guidelines; for example, only women aged ≥40 years are included in the denominator for mammography. Colonoscopy was considered completed if performed within 2 years of sample collection, whereas all other USPSTF-recommended screenings were required to be completed within 6 months of sample collection.
[B]Additional Cancer Screenings: All additional screening tests were required to be performed within 6 months before or any time after dermatomyositis (DM) symptom onset. N/A = no patients were eligible.

**Supplemental Table 7. Serial ctDNA Measurement**

| Systemic Sclerosis | | | Dermatomyositis | | |
|---|---|---|---|---|---|
| | **Available 2nd sample** | **2nd sample positive** | | **Available 2nd sample** | **2nd sample positive** |
| **Baseline ctDNA positive (N=11)** | 5 of 11 (45%) | 0/5 (0%) | **Baseline ctDNA positive (N=6)** | 6 of 6 (100%) | 2 of 6 (33%)[A] |
| **Baseline ctDNA negative (N=58)** | 22 of 58 (38%) | 0/22 (0%) | **Baseline ctDNA negative (N=59)** | 40 of 59 (68%) | 3 of 40 (8%)[B] |

[A]Of these 2 patients, one was diagnosed with cancer (MYOS59), whereas one remained cancer-free. [B]Three DM patients had a positive result on their second ctDNA test (MYOS43, MYOS47, and MYOS112). MYOS43 was diagnosed with cancer 1.4 months after the positive ctDNA result, and the same mutation was identified in the tumor. In contrast, MYOS47 and MYOS112 remained cancer-free.

**Supplemental Table 8: Rheumatic Disease Center-Specific Approach to Newly Diagnosed Patients**

| Cancer Screening Test Recommended | Scleroderma Center | Myositis Center |
|---|---|---|
| **Female Only** | | |
| *Mammogram per USPSTF Guidelines* | X | X |
| *Transvaginal Ultrasound* | | X |
| *Pap Smear per USPSTF Guidelines* | X | X |
| **Male Only** | | |
| *PSA* | X | X |
| **Female and Male** | | |
| *Complete Blood Count* | X | X |
| *Complete Metabolic Panel* | X | X |
| *Serum Protein Electrophoresis* | X | X |
| *High-resolution CT chest* | X | |
| *CT chest/abdomen/pelvis with IV contrast OR PET/CT*[A] | | X |
| *Colonoscopy per USTSPF Guidelines* | X | X |

Age-appropriate cancer screening per U.S. Preventive Services Task Force (USPSTF) recommendations included mammography (≥40 years), Papanicolaou smear (≥21 years), colonoscopy (≥45 years as of 2021), and prostate-specific antigen testing with shared decision-making for individuals aged 55–69 years. Chest CT was frequently obtained for evaluation of interstitial lung disease; [A]patients positive for RNA polymerase III may have participated in a separate research imaging protocol involving whole-body PET/CT.
